# PRELIMINARY ASSESSMENT OF STRUCTURE AND COLLECTIVE DOSE FROM CT EXAMINATIONS RELATED TO COVID-19 DIAGNOSTICS IN THE RUSSIAN FEDERATION IN MARCH - JUNE 2020

**DOI:** 10.1101/2020.08.25.20181396

**Authors:** A.V. Vodovatov, I.K. Romanovich, O.A. Istorik, L.A. Eremina, S.P. Morozov, S.A. Ryzhov, G.V. Berkovich, I.G. Kamyshanskaya, G.E. Trufanov, L.A. Chipiga, P. S. Druzhinina, A.M. Biblin, R.R. Akhmatdinov, I.V. Basek, A.A. Karatetskiy, D. V. Merkulov, Yu.S. Ispravnikova, V.V. Pritz, N.S. Polishchuk, A.N Mukhortova, V.G. Pyzyrev

## Abstract

The use of computed tomography (CT) for the diagnostics of COVID-19 in the Russian Federation led to significant changes in the structure of X-ray diagnostics and levels of medical exposure of the patients. This study was aimed at the preliminary operative assessment of changes in the structure and collective dose from CT examinations in several representative hospitals, regions and on the level of the Russian Federation. The results of the study indicate that during the transformation of hospitals from general medical practice into dedicated COVID-19 facilities, the number of CT examinations increased up to 30%; the collective dose from CT exams increased up to a factor of 1.5. During a partial transformation of a medical facility into the hospital with separate COVID-19 departments, the increase in the number of CT examinations in the facility was more significant (up to a factor of 2 or more). These numbers correspond to 1.5 - 2.5 chest CT examinations (from 1 to 6) per patient admitted to hospital with COVID-19 diagnosis; and 1.2 chest CT examinations per patient in outpatient facilities, including a mandatory CT scan for the staging of COVID-19. The collective dose from CT examinations in the Russian Federation for March-June period of 2020 increased by the factor of 2 (from 16k man-Sv to 32k man-Sv); the collective dose of COVID-19 patients was about 12k man-Sv. For a more detailed and reliable assessment of the dynamics of changes in the structure of diagnostic radiology and levels of radiation exposure of patients in the Russian Federation, data collection in the regions of the Russian Federation and individual medical facilities will continue.

## Introduction

The novel coronavirus infection (COVID-19) is an infectious disease caused by SARS-CoV-2 virus [1]. At the current moment, the virus has spread to almost all countries of the world. As of July 17, 2020, the coronavirus infection was detected in more than 13.5 million of people; more than 584 thousand people died already [1, 2]. In the Russian Federation, these numbers correspond to about 759 thousand and 12 thousand people respectively [3].

The use of computed tomography (CT) for the diagnostics of COVID-19 has been widely discussed within the medical community. Initially, there were several points of view on the applicability of diagnostic imaging modalities, ranging from the use of CT scan for screening of the disease [4] to the use of CT only for confirmed cases of COVID-19 [5]. This discussion is related to the requirement to understand the diagnostic significance of the CT study, as well as the availability of healthcare resources and this particular method in regions or in the country, and to the current epidemiological situation.

Due to the fact that reliability of PCR-method of diagnostics did not exceed 70% in many countries, including Russia, a decision was made to introduce the concept of “clinically confirmed case of COVID-19”, which combines all symptoms, the presence of respiratory disorders and results of CT-or radiography examinations (regardless of a single PCR test result for the presence of SARS-CoV-2 RNA and epidemiological anamnesis) [6]. The methods of diagnostic radiology were used not only to identify COVID-19, pneumonia, complications, or for differential diagnosis with other lung diseases, but to determine the severity and dynamics of changes, as well as to assess the effectiveness of therapy [7]. Additionally, it should be noted that CT scan is not recommended and should not be used as a screening method for the diagnosis of coronavirus infection.

However, due to the availability, high diagnostic informativity, non-invasiveness and fast speed, CT scan becomes the indispensable diagnostic method for the early and initial diagnosis of COVID-19. CT scan allows suspecting the diagnosis of COVID-19 viral pneumonia, quickly assess the extent of lung tissue damage and its severity, and to clarify the stage of changes according to the characteristics of novel coronavirus pneumonia. Based on CT scan results and clinical data, as well as on patient’s history, it becomes possible to quickly route patients and initiate antiviral therapy [7,8].

In the Russian Federation, changes in the structure of diagnostic radiology associated with COVID-19 would lead to changes in the level and structure of the collective dose to the public from medical exposure. This process is bidirectional. On one hand, the number of chest CT scans significantly increases. On the other hand, a transformation of some hospitals exclusively for treatment of patients with COVID-19, the closure of some medical facilities or departments due to COVID-19 quarantine, would lead to decrease of scheduled CT exams. Such CT exams primarily include multiphase contrast-enhanced CT scans, associated with high (up to 50-80 mSv) individual patient doses [9, 10]. Unfortunately, at the current moment, there is no reliable data on changes of the diagnostic radiology structure and exposure levels of the Russian population due to COVID-19 pandemic.

The purpose of this study was to perform a preliminary prognostic assessment of changes in the collective dose of the population of the Russian Federation from CT exams during the COVID-19 pandemic (March – June 2020). The available data trends of CT exams were analyzed for several hospitals in different regions of the Russian Federation.

## Materials and methods

Primary data was collected in selected hospitals of St. Petersburg (Almazov National Medical Research Centre, Urban Mariinsky Hospital), that had been transformed into COVID-19 hospitals in April - May 2020. The data was collected by analyzing patient medical records and data from the radiological information system (PACS). The following values were assessed for the period of February – May 2020: the total number of CT exams performed in the hospital and the number of CT scans performed on patients hospitalized with COVID-19. Additionally, the number of CT exams per patient for the entire period of his/her hospital stay was estimated.

Data on CT scans performed for COVID-19 diagnostics in some regions of the Russian Federation (Moscow, Leningrad region), were obtained from the regional healthcare departments. The number of CT exams was estimated in both outpatient facilities and hospitals.

Predictive assessment of changes in the number of CT scans and in collective dose from CT exams in the Russian Federation was performed by analyzing the regional data from the national dose data collection system (3-DOZ^1^ ^2^ form with 2019 data) according to the following scheme:

–estimation of the number of different types of CT exams for 1 month in each region according to the equation (1). For the conservative estimation it was assumed that the number of CT examinations in the region for 1 month during the COVID-19 pandemic in 2020 has not changed compared to 2019.

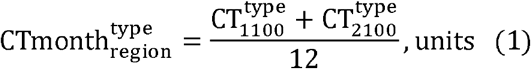

where: 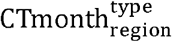 - the number of CT exams of the selected anatomical area in the selected region for 1 month; 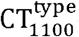 - the number of CT exams of the selected anatomical area from column 8 of table 1100 from the 3-DOZ form (the number of examinations with calculated patient doses); 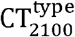 - the number of CT exams of the selected anatomical area from column 8 of table 2100 from the 3-DOZ form (the number of examinations with measured patient doses);

– estimation of the number of different types of CT examinations for 4 months (March - June 2020) during COVID-19 pandemic in each region according to the equation (2).

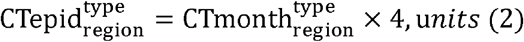

where: 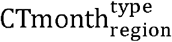 - the number of CT exams of the selected anatomical area in the selected region for 1 month; 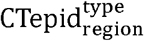 - the number of CT exams of the selected anatomical area in the selected region for 4 months.

– assessment of average individual effective doses for CT examinations in the region according to the equation (3):

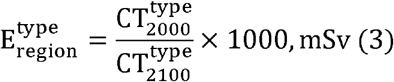

where: 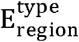 - the average individual effective dose for CT exam of the selected anatomical area in the selected region, mSv; 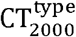 – the collective dose from CT exam of the selected anatomical area in the selected region, man-Sv; 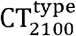 – the number of CT exams of the selected anatomical area from column 8 of table 2100 from the 3-DOZ form (the number of examinations with measured patient doses).

–assessment of changes in the number of CT examinations and in the collective dose from CT scans.

–estimation of the collective dose from CT examinations according to the equation (4):

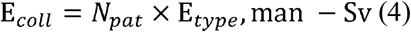

where: E*_coll_* - the collective dose from CT exams, man - Sv; *N_pat_* - the number of patients who underwent the selected CT exams, people; *E_type_* - the average individual effective dose from CT exams of the selected anatomical area in the selected region, mSv.

– assessment of the risk of radiation-induced malignancies and hereditary diseases from CT examinations per 1 million people in the Russian Federation according to the equation (5):

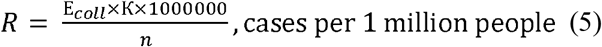

where: R - the number of possible cases of radiation-induced malignant neoplasms and hereditary diseases from CT exams per 1 million people in the Russian Federation, cases per 1 million people; E*_coll_* - collective dose from CT exams, man-Sv; K – the linear coefficient of radiation risk - 5,7 × 10 ^-2^ Sv ^-1^^3^; n - the Russian population (146 700 000 people).

Statistical data processing (descriptive statistics for samples, comparison of samples using nonparametric statistics methods) was performed using Statistica 10 Software.

## Results and discussion

### 1. Data from the selected medical facilities

Table 1 presents the data on the number of CT examinations performed for COVID-19 patients in Almazov National Medical Research Centre (NMRC), St. Petersburg, Russia. Data are presented for a sample of 167 patients admitted to the hospital in May 2020.

**Table 1.** The number of CT examinations per patient for the hospital stay in NMRC, depending on the disease severity. * data are presented in the format: mean ± standard deviation (minimum-maximum)

| Diagnosis/Severity classification | CT-0 | CT-1 | CT-2 | CT-3 | CT-4 |
| --- | --- | --- | --- | --- | --- |
| Lung tissue damage | 0 | <25% | 25-50% | 50-75% | > 75% |
| Number of patients | 12 | 19 | 46 | 47 | 14 |
| Number of CT scans per patient per hospital stay* | 2.3 ± 1.1<br>(1-4) | 2.4 ± 1.2<br>(1-6) | 2.3 ± 1.0<br>(1-5) | 2.7 ± 1.3<br>(1-5) | 2.4 ± 1.1<br>(1-5) |
| Number of patients who were underwent more than 3 CT scans | 2 | 2 | 7 | 13 | 2 |
| Average number of CT scans per patient | 2.5 |  |  |  |  |

<sup>3</sup> Sanitary norms and rules 2.6.1.2523-09. Norms of the radiation safety.
\* data are presented in the format: mean ± standard deviation (minimum-maximum)

No significant differences were indicated in the number of CT examinations performed per patient with different stages of COVID-19 (pairwise comparison using the Mann-Whitney test, p> 0.05). 2.5 CT scans were performed per patient on average (1 CT exam during admission and 1-2 CT scans to monitor a disease dynamics). However, only 26 patients (20% from the whole sample) received more than 3 CT exams. Analysis of patient records in St. Petersburg Urban Mariinsky Hospital indicated that COVID-19 patients received 1.4 CT exams per patient on average (3656 scans for 2607 patients and 3400 scans for 2386 patients in May and June respectively). These numbers correspond to the recommended frequency of CT examinations by the COVID-19 clinical guidelines, published by the Ministry of Health of the Russian Federation [11, 12]. It should be noted, that the decision on the number of control CT examinations is made by the referring physician based on the clinical indications [11, 12].

Data trends of the structure and the collective dose from CT examinations in NMRC is presented in tables 2 and 3, respectively.

**Table 2.** The structure of CT - examinations in NMRC for some months of 2020.

| Parameter | February |  | May |  | June |  |
| --- | --- | --- | --- | --- | --- | --- |
|  | without contrast | with contrast | without contrast | with contrast | without contrast | with contrast |
| Number of CT exams | 1898 | 919 | 1509 | 459 | 1167 |  |
| Total | 2817 |  | 1969 |  | 1167 |  |
| Change in the number of CT exams relative to February 2020, % | - |  | -30% |  | -58% |  |
| Number of chest CT exams | 820 | 429 | 969 | 215 | 1167 |  |
| Total | 1249 |  | 1185 |  | 1167 |  |
| Change in the number of chest CT exams relative to February 2020, % | - |  | -5% |  | -7% |  |

**Table 3.** The structure of the collective dose from CT examinations in NMRC for some months of 2020.

| Type of CT examination | February |  | May |  | June |  |
| --- | --- | --- | --- | --- | --- | --- |
|  | CT without contrast | CT with contrast | CT without contrast | CT with contrast | CT without contrast | CT with contrast |
| Collective Dose from CT exams, man-Sv | 12.1 | 13.9 | 9.7 | 6.9 | 7.6 | - |
| Total, man-Sv | 26.0 |  | 16.6 |  | 7.6 |  |
| Change in the collective dose relative to February 2020, % | - |  | -36% |  | -71% |  |
| Collective Dose from chest CT exams, man-Sv | 5.3 | 6.2 | 6,3 | 3.1 | 7.6 | - |
| Total, man-Sv | 11.5 |  | 9.4 |  | 7.6 |  |
| Change in the collective dose from chest CT exams relative to February 2020, % | - |  | -18% |  | -34% |  |

Data trends of the structure and the collective dose from CT examinations in St. Petersburg Urban Mariinsky Hospital is presented in tables 4 and 5, respectively.

**Table 4.** The structure of CT examinations in St. Petersburg Urban Mariinsky Hospital in February-June 2020.

| Month | February |  | March |  | April |  | May |  | June |  |
| --- | --- | --- | --- | --- | --- | --- | --- | --- | --- | --- |
| Type of CT examination | without contrast | with contrast | without contrast | with contrast | without contrast | with contrast | without contrast | with contrast | without contrast | with contrast |
| Number of exams | 2716 | 136 | 2580 | 136 | 2535 | 54 | 3747 | 4 | 3660 | ten |
| Total | 2852 |  | 2716 |  | 2589 |  | 3751 |  | 3670 |  |
| Change in the number of CT exams relative to February 2020, % | - |  | -5% |  | -9% |  | + 32% |  | + 29% |  |
| Number of chest CT exams | 458 |  | 485 |  | 1232 |  | 3342 |  | 3155 |  |
| Change in the number of chest CT exams relative to February 2020, % |  |  | + 6% |  | + 169% |  | + 630% |  | + 589% |  |

**Table 5.** The structure of the collective dose from CT examinations in St. Petersburg Urban Mariinsky Hospital in February-June 2020.

| Month | February |  | March |  | April |  | May |  | June |  |
| --- | --- | --- | --- | --- | --- | --- | --- | --- | --- | --- |
| Type of the CT examination | without contrast | with contrast | without contrast | with contrast | without contrast | with contrast | without contrast | with contrast | without contrast | with contrast |
| Collective Dose from the CT exams, man-Sv | 14,0 | 2 | 14.3 | 2 | 14.2 | 0.7 | 23.5 | 0.1 | 23 | 0.2 |
| Total, man-Sv | 16,0 |  | 16.3 |  | 14.9 |  | 23.6 |  | 23.2 |  |
| Change in the collective dose relative to February 2020, % | - |  | + 2% |  | -7% |  | + 47% |  | + 45% |  |
| Collective Dose from chest CT exams, man-Sv | 4.2 |  | 4.5 |  | 8.7 |  | 21.9 |  | 20.6 |  |
| Change in the collective dose from chest CT exams relative to February 2020, % | - |  | + 7% |  | + 105% |  | + 415% |  | + 387% |  |

According to the tables 2 and 3, the collective dose from CT examinations rapidly decreased in NMRC: from 26 man-Sv in February to 16.6 man-Sv in May and 7.6 man-Sv in June (up to three times lower). It can be explained by a slight increase in the number of chest CT exams, and at the same time, a rapid decrease in the number of all other CT examinations (including highdose CT scans with contrast). It should be noted, since transforming NMRC into the Covid-19 center (May 15, 2020), only chest CT scans have been performed (560 in May, 1167 in June).

On the contrary, in St. Petersburg Urban Mariinsky Hospital, the number of CT examinations increased by 30% from February to June, 2020. At the same time, in June, the number of chest CT scans increased by a factor of 7 with almost total absence of other CT examinations. The changes in the number and structure of CT examinations led to 45% increase in the collective dose from CT studies (from 16 to 23 man - Sv). The main increase in the number of chest CT scans and a collective dose corresponds to the moment of transforming Urban Mariinsky Hospital into the COVID-19 hospital.

### 2. Analysis of data from some regions of the Russian Federation

Data on the number of chest CT examinations is presented in Tables 6 and 7 for the Leningrad region and city of Moscow, respectively.

**Table 6.** Changes in the number of CT examinations and the corresponding collective dose in the Leningrad Region. Data are presented for the first 6 months of 2019 and 2020 (January - June period)

| Year | 2019 |  | 2020 |  |
| --- | --- | --- | --- | --- |
| Examinations | Total | Chest | Total | Chest |
| Number of CT exams | 85997 | 19586 | 92656 | 36865 |
| Change in the number of CT exams relative to 2019, % |  |  | +8% | +88% |
| Collective dose from CT exams, man-Sv |  | 84,2 |  | 158,5 |
| Change in the collective dose relative to 2019, % | +88% |  |  |  |

**Table 7.** Changes in the number of CT examinations and the collective dose for the period from March to June 2020 in Moscow.

| Parameter | Type of medical facility | March | April | May | June |
| --- | --- | --- | --- | --- | --- |
| Number of chest CT exams | Outpatient clinics | 6247 | 49 317 | 94614 | 40414 |
|  | Hospitals | 3 391 | 9,230 | 21858 | 22719 |
| Total |  | 9 638 | 58 547 | 116 472 | 63,133 |
| Change in the number of chest CT exams relative to 2019, % |  | -78% | + 35% | + 168% | + 45% |
| The number of CT-examinations per patient in outpatient clinics |  | 1.0 | 1.1 | 1.3 | 1,2 |
| Collective dose from chest CT exams, man-Sv | Outpatient clinics | 30 | 236.7 | 454.1 | 194.0 |
|  | Hospitals | 16.3 | 44.3 | 104.9 | 109.0 |
| Total |  | 46.3 | 281.0 | 559.1 | 303, 0 |
| Change in the collective dose relative to 2019, % |  | -78% | + 35% | + 168% | + 45% |

According to Table 6, the number of chest CT scans in the Leningrad Region and the corresponding collective dose increased by 2 times in 2020. At the same time, the total number of CT exams in this region did not significantly change (increased by less than 10%). Most hospitals reported increase in the number of chest CT examinations by a factor of 1.5-8 (2 on average) compared to the first six months of 2019 due to decrease in the number of CT exams of all other anatomical areas (by 50% on average). Unfortunately, reliable estimation of changes in the collective dose from CT examinations is not possible due to the lack of data on the structure of CT diagnostics in Leningrad region.

Table 7 presents the data on the number of chest CT examinations performed in Moscow medical facilities for the period of March - June 2020.

The data from table 7 indicates systematic increase in the number of chest CT examinations in both outpatient and inpatient facilities. A maximum number of CT exams – 116,000 - was performed in May 2020, which is higher by a factor of 2.8 compared to 2019. Low number of CT examinations in March is explained by the limited data collection in that month. The number of CT studies in April - June 2020 corresponds to the data collected from 80% of medical facilities in Moscow. The dynamics of the collective dose from chest CT examinations correlates with the dynamics of the number of CT exams – the maximum increase in the collective dose by a factor of 3 compared to 2019 was recorded in May. In the remaining months, the increase in the collective dose from chest CT scans did not exceed a factor of 1.5 compared to 2019.

It should also be noted that an average of the number of CT examinations per patient with suspected COVID-19 in outpatient setting was significantly lower compared to the data from St-Petersburg - 1.2 CT exams per patient.

It is important to note the features of how diagnostic radiology is organized in Moscow. To reduce the burden on the healthcare system, the majority of CT examinations were performed at the prehospital stage, i.e. in outpatient facilities [6, 13-15], backed up with the extensive use of telemedicine technologies and establishing reference centers for diagnostic radiology services [16]. This resulted in a significant reduction in the number of hospitalizations and hospitals’ workload and in optimization of healthcare resources. Apparently, it also led to a lower number of CT examinations per patient.

At the same time, the application of the best practices in radiology, widespread use of CT, and the need for dynamic patient monitoring with repeated CT scans [17] require detailed consideration of radiation safety and long-term radiation risks both for individual patients and on the population level.

The following features of data on CT examinations in regions of the Russian Federation are the following:

-The lack of detailed data on the structure of CT examinations of other anatomical areas (including contrast-enchanced examinations);

-The lack of detailed data on changes in the structure of other radiological and nuclear medicine examinations;

-The absence of data on typical patient doses (average effective dose) from CT examinations as well as from other radiological and nuclear medicine studies.

### 3. The assessment of changes in the collective dose of the population of the Russian Federation from CT examinations

The assessment of the change in the collective dose of the population of the Russian Federation for four months of 2020 (March - June) related to diagnosis of COVID-19 is presented in Table 8 for the following scenarios:

**Table 8.** The changes in the collective dose of the population of the Russian Federation and the corresponding number of radiation-induced cancers and hereditary effects for March - June 2020 for various scenarios.

| Parameters | 3-DOZ | Scenarios |  |  |  |  |  |
| --- | --- | --- | --- | --- | --- | --- | --- |
|  | 4 months 2019 | Covid-19 hospital |  | Intense activity |  | Combined option |  |
|  | Collective dose, man - Sv | Collective dose, man- Sv | % * | Collective dose, man - Sv | % | Collective dose, man - Sv | % |
| Chest CT exam | 4836 | 17917 | + 270% | 21187 | + 338% | 18734 | + 287% |
| CT exam of other anatomical areas | 11239 | 2248 | -80% | 11239 | 0% | 4496 | -60% |
| Total | 16075 | 20164 | + 25% | 32426 | + 102% | 23230 | + 45% |
| Cases of radiation-induced cancers and hereditary effects, units | 916 | 1149 |  | 1848 |  | 1324 |  |
| Exceedance compared to 2019 |  | 233 | + 25% | 932 | + 102% | 408 | + 45% |
| Cases per 1 million population |  | 2 |  | 6 |  | 3 |  |

\* change in the collective dose for the period of March - June 2020 relative to the same period in 2019, %

***Scenario 1. “Covid-19hospital”***. It corresponds to the transformation of general practice regional hospitals into hospitals for COVID-19 patients and emergency patients with various pathologies. In this scenario, we assume that the total number of CT examinations in the region has not been changed; the number of CT exams of all other anatomical areas (except for the chest) decreased by 80%. The released capacities were used for chest CT scans. This scenario is based on results of the data analysis of medical facilities in St. Petersburg (Tables 2-5).

***Scenario 2. “Intense activity”***. Corresponds to the medical facilities operating after the peak of COVID-19: the number of examinations remains the same as in 2019, with the exception of chest CT scan. We assume that prior to each admission to the hospital or planned X-ray examination, a patient undergoes chest CT examination. The number of CT studies increases by the sum of all other CT exams. This scenario is extremely conservative and is based on the analysis of data from the regions of the Russian Federation.

***Scenario 3. “Combined”***. A combination of scenario “Covid-19 hospital” for 3 months with scenario “Intensive activity” for 1 month.

According to the data from Table 8, increase in the collective dose from CT examinations for the period of March - June 2020 compared to the same period in 2019 does not exceed a factor of 2 for the “intensive activity” scenario. “Combined” scenario looks more real, the collective dose from CT examinations increases by 50%. For the worst scenario, the number of radiation-induced cancers and hereditary effects, associated with CT examinations does not exceed 6 cases per 1 million of the population.

It is possible to conservatively estimate the collective dose and the associated additional population radiation risk for COVID-19 patients. During the COVID-19 pandemic in the Russian Federation as of July 18, 2020, 759,000 cases of COVID-19 were detected. If we assume that all people with COVID-19, regardless of hospitalization, underwent 2.5 chest CT exams (see Table 1) with an average dose of 6.5 mSv per examination [9, 10], the collective dose for this cohort corresponds to 12,334 man-Sv, which, in turn, corresponds to 703 additional cases of radiation-induced cancers and hereditary effects associated with CT diagnostics for this cohort, or 5 cases per 1 million of the population of the Russian Federation.

These presented scenarios allow evaluating changes in the collective dose only from CT examinations and do not consider the changes in the structure of X-ray and nuclear medicine diagnostics in the Russian Federation during the COVID-19 pandemic:

- decrease in the number of planned hospital admissions and associated decrease in the number of high-dose examinations (multiphase contrast-enhanced CT scans, interventional radiology, hybrid nuclear medicine examinations, etc.);
- increase in the number of conventional radiography of the chest performed for the patients in intensive care units or when CT scan is not available;
- decrease in the number of X-ray examinations of all other anatomical areas;
- decrease in the number of dental and screening examinations (fluorography, mammography).

Unfortunately, now it is not possible to perform a comprehensive assessment of the change of the collective dose from medical exposure in the Russian Federation due to the lack of primary data on changes in the structure of all types of diagnostic radiology exams during the COVID-19 pandemic.

## Conclusions

COVID-19 pandemic in the Russian Federation had a significant impact on the structure of diagnostic radiology and was associated with changes in the collective dose in different regions of the Russian Federation. This study presents the first conservative quantitative estimates of changes in collective doses from CT examinations used for COVID-19 diagnosis on different levels: from the single medical facility to the country level.

First of all, significant drawbacks of the existing radiological data collection systems were identified. At the time of this writing, the detailed data on the number of performed CT examinations (both for COVID-19 diagnosis and all other reasons) is available only at the level of a medical facility. The centralized data collection on the regional level was not performed and/or available. The exception is Moscow, where all outpatient facilities and about 80% of hospitals are integrated into a joint medical electronic information system [18]. However, even in the presence of the unified medical electronic information system, data on patient doses are not automatically collected so far.

The federal statistical reporting forms (radiation-hygienic passport [4], 3-DOZ form of the ESKID system, Form-30 of the Ministry of Health of the Russian Federation [5]) are submitted once per year. Hence, it is necessary to upgrade the existing federal data reporting forms to promptly provide up-to-date information of dynamics of processes in diagnostic radiology and to introduce automated electronic patient dose data collection. Data on individual patient doses is available only at the level of the individual medical facility, reducing the reliability of collective dose estimations.

Data obtained from individual medical facilities and regions of the Russian Federation allow us to draw the following conclusions:

- on average, 1.5 - 2.5 chest CT examinations (from 1 to 6) were performed per patient admitted to hospital with COVID-19 diagnosis; and 1.2 chest CT examinations per patient in outpatient facilities. These indicators include mandatory chest CT scans during hospitalization. A physician makes a decision on the frequency and number of control CT exams.
- during the transformation of hospitals from general medical practice into dedicated COVID-19 facilities, the number of CT examinations increased up to 30%; the collective dose from CT exams increased up to a factor of 1.5. This is explained by a rapid (up to a factor of 3) increase in the number of chest CT exams and a decrease in the number of CT scans of other anatomical areas.
- during a partial transformation of a medical facility into the hospital with separate COVID-19 departments, the increase in the number of CT examinations in the facility was more significant (up to a factor of 2 or more). This can be explained by performing chest CT scans for COVID-19 patients in combination with all other CT exams carried out according to the treatment protocol of patients with other diseases, as well as chest CT-examinations for everyone admitted to the hospital to exclude COVID-19;
- in the regions of the Russian Federation where the study was conducted, the collective dose from CT examinations increased by the factor of 2 (up to a factor of 3 in some months in the city of Moscow).
- the lack of data on changes in the structure of diagnostic radiology in the regions of the Russian Federation does not allow assessing changes in the entire collective dose from medical exposure.

Assessment of the dynamics of the collective dose of the population of the Russian Federation from CT studies in the period from March to June 2020 allows us to draw the following conclusions:

- the collective dose from CT examinations in the Russian Federation for four months of 2020 increased by the factor of 2 (from 16k man-Sv to 32k man-Sv) corresponding to the scenario “intense activity”;
- the collective dose of COVID-19 patients, based on 2.5 CT examinations per patient, was about 12k man-Sv;
- this increase in collective dose would be associated with additional 5-6 cases of radiation-induced cancers and hereditary effects in addition to the background morbidity per 1 million population of the Russian Federation.

For a more detailed and reliable assessment of the dynamics of changes in the structure of diagnostic radiology and levels of radiation exposure of patients in the Russian Federation, it is necessary to collect data in the regions of the Russian Federation and individual medical facilities.

## Data Availability

All the data usied in the preparation of the manuscript is available by request from the authors.

## Footnotes

1 Order of the Ministry of Healthcare of the Russian Federation #298, 31.07.2000 “On the establishment of the joint governmental system of the control and registration of the individual doses of the citizens"

2 Rospotrebnadzor. Methodical recommendations.Ni) 0100/1659-07-26 “Completion of the federal governmental statistical surveillance form 3-DOZ”

3 Sanitary norms and rules 2.6.1.2523-09. Norms of the radiation safety.

